# Diarrheagenic Escherichia coli in children under five in the Americas: Prevalence, pathotype distribution, and socioenvironmental drivers—A systematic review

**DOI:** 10.1101/2025.06.19.25329931

**Authors:** Diego Fernando López Muñoz, Beatriz Giraldo Ospina, Cristhian Camilo Velandia Mosquera, Leonardo Beltrán Angarita, Yamil Liscano Martínez, Vanessa Muñoz Valencia

## Abstract

**Aim:** To characterize the prevalence, pathotype distribution, and socioenvironmental drivers of diarrheagenic *Escherichia coli* (DEC) in children under five in the Americas.

**Methods:** A systematic review and meta-analysis of studies (2015-2024) from PubMed, Scopus, Springer, SciELO, LILACS and Web of Science were conducted. Eligible studies reported DEC prevalence in children ≤ 5 years with acute diarrhea using molecular/microbiological diagnostics.

**Results:** Thirteen studies (7,485 participants) were included. Pooled DEC prevalence was 28.3% (95% CI: 21.6-35.7). PCR detected 1.4-fold higher prevalence than culture. Regional disparities included EPEC dominance in the Andean highlands (50.0%) and EAEC/EHEC co-occurrence in Amazonia (EAEC 8x post-hurricane). Limited water access (aOR 3.2) and poor sanitation were key risk factors; a 10% increase in water access reduced EPEC burden by 17%.

**Conclusion:** DEC poses a significant health burden in the Americas, with pathotype distribution linked to socioeconomic inequities. Molecular diagnostics are crucial for surveillance, and policy should prioritize WASH infrastructure and equitable diagnostics access to mitigate morbidity, antimicrobial resistance, and climate-amplified outbreaks.

**Summary:**

- Regional variations in the prevalence of diarrhoeagenic *Escherichia coli* and associated risk factors in children under five in the Americas were poorly understood, necessitating a systematic review.
- The study found a prevalence of 28.3%, with regional disparities and socioeconomic factors such as limited access to water driving infections.
- Improved water, sanitation and hygiene infrastructure, as well as molecular diagnostics, are critical for reducing paediatric diarrhoeal morbidity and antimicrobial resistance.

## 1. Background

Diarrhoeal diseases pose a significant public health challenge in the Americas, with a disproportionate impact on children under five [1]. Despite the global efforts that have been made to reduce under-five mortality, diarrhoeagenic *Escherichia coli* (DEC) remains a significant yet frequently underdiagnosed cause of paediatric illness [2]. The DEC has been identified as a causative agent in approximately 30% of acute diarrhoea cases in this age group within the region [3]. The six primary DEC pathotypes — enteropathogenic (EPEC), enterotoxigenic (ETEC), enteroaggregative (EAEC), enterohemorrhagic (EHEC), enteroinvasive (EIEC) and diffusely adherent (DAEC) — possess distinct virulence characteristics and ecological niches [4]. This phenomenon gives rise to a diverse array of disease burdens across a range of geographical, socioeconomic and climatic landscapes [2,3].

A number of critical knowledge gaps have been identified as factors that hinder effective control of DEC in children [5]. A significant concern pertains to the persistent utilisation of conventional culture-based diagnostics in numerous clinical laboratories, particularly in Low-Middle Income Countries (LMICs), which results in a 20–40% underestimation of DEC prevalence when compared with Polymerase Chain Reaction (PCR) methods [6]. This has the effect of obscuring the true burden of pathotypes and delaying outbreak detection. Furthermore, the epidemiology of DEC varies considerably between different geographical regions, and this variation is not yet fully understood [2]. For instance, EPEC predominates in the rural Andean highlands, driven by inadequate water supplies, while the Amazon experiences co-circulation of EAEC and EHEC, with EAEC surging after extreme weather events, as evidenced in Honduras in 2022 [7]. These patterns are indicative of complex environmental, social and microbial dynamics. However, detailed local data are limited.

The challenge of controlling DEC is exacerbated by the limited monitoring of antimicrobial resistance (AMR), with up to 35% of isolates demonstrating multidrug resistance, a phenomenon that is intensified by the inappropriate use of antibiotics in 35% of paediatric diarrhoea cases [8–10]. The repercussions of the pandemic have precipitated a resurgence in the prevalence of DEC in marginalised communities, with disruptions to healthcare and vaccination programmes serving as key catalysts. It is evident that climate change exerts a significant influence on the DEC patterns, yet the development of predictive models that incorporate climate risk into health planning remains in its infancy [11]. It is imperative that these gaps are addressed in order to facilitate effective communication and collaboration. The incorporation of PCR into routine surveillance procedures has been demonstrated to enhance detection accuracy, thereby generating a return on investment of $6.20 for every dollar invested, through the reduction of antimicrobial resistance-related hospitalisation [12]. Furthermore, the implementation of WASH interventions is imperative. For instance, a 10% increase in water access has been shown to result in a 17% reduction in EPEC burden [13–15].

The present systematic review has been conducted with the objective of synthesising recent data in order to quantify the prevalence of DEC, map the distribution of its pathotypes, and identify the key socioeconomic and environmental determinants in children under five across the Americas. By comparing molecular and conventional diagnostic methods, this review also highlights methodological biases and proposes a framework for more precise public health interventions. The aim of the present study is to provide the scientific community with findings that can inform evidence-based policies. Such policies are intended to reduce the burden of DEC, mitigate AMR, and promote child health equity in a region facing ongoing and emerging enteric disease threats.

## 2. Methods

### 2.1 Study design and registration

This systematic review and meta-analysis adhered to PRISMA 2020 guidelines [16] and was prospectively registered with PROSPERO (CRD420251059483) [17]. We included observational studies (cross-sectional, cohort, or surveillance-based) reporting prevalence or incidence of diarrheagenic *Escherichia coli* (DEC) in children aged ≤5 years with acute diarrhea across the Americas, diagnosed via microbiological or molecular methods.

### 2.2 Search strategy and selection criteria

A comprehensive search was conducted across six databases (PubMed, Scopus, Web of Science, SciELO, LILACS, Springer) for studies published between January 2015 and December 2024. The search strategy combined controlled vocabulary and free-text terms:

(“diarrheagenic” OR “pathogenic” OR “enteropathogenic” OR “enterotoxigenic” OR “enteroaggregative” OR “enterohemorrhagic” OR “enteroinvasive” OR “diffusely adherent”) AND “Escherichia coli” AND (“prevalence” OR “incidence” OR “occurrence” OR “frequency”) AND (“pediatric” OR “children” OR “child” OR “infant”) AND (“Americas” OR “North America” OR “South America” OR “Latin America” OR “Central America” OR “Caribbean”).

### 2.3 Eligibility criteria

To ensure the systematic review included relevant and high-quality studies, we applied strict inclusion and exclusion criteria guided by the research question and objectives, focusing on diarrheagenic *Escherichia coli* (DEC) epidemiology in children under five in the Americas. These criteria are detailed in Table 1.

**Table 1.** Inclusion and exclusion criteria for systematic review of diarrheagenic *Escherichia coli* (DEC) in children under five in the Americas.

| **Inclusion criteria** |  |  |
| --- | --- | --- |
| **Criterion type** | **Criterion** | **Rationale** |
| Study population | Original studies reporting primary data on DEC prevalence or incidence in children aged $\leq 5$ years with acute diarrhea. | Ensures focus on pediatric populations most vulnerable to DEC, aligning with the review's objective to quantify disease burden in this age group. |
| Geographic scope | Studies conducted in the Americas (North, Central, South America, or Caribbean). | Restricts analysis to the target region to address regional epidemiology and disparities, as specified in the study aims. |
| Diagnostic methods | Studies using microbiological (e.g., culture, ELISA) or molecular (e.g., PCR, metagenomics) diagnostic methods to identify DEC pathotypes (EPEC, ETEC, EAEC, EHEC, EIEC, DAEC). | Ensures diagnostic rigor and allows comparison of method-specific detection rates, addressing underdiagnosis due to low-sensitivity techniques. |
| Publication language | Studies published in English, Spanish, or Portuguese. | Reflects predominant languages of scientific literature in the Americas, maximizing inclusion of relevant studies while maintaining accessibility for review team |
| Study design | Observational studies (cross-sectional, cohort, or surveillance-based) providing prevalence or incidence data. | Focuses on study designs suitable for epidemiological estimates, ensuring robust data for meta-analysis and regional comparisons. |
| Publication period | Studies published between January 2015 and December 2024. | Captures recent data to reflect current DEC epidemiology, including post-COVID-19 trends and advancements in diagnostic technologies. |
| **Exclusion criteria** |  |  |
| **Criterion type** | **Criterion** | **Rationale** |
| Study type | Case reports, case series, narrative reviews, or editorials. | Excluded to prioritize studies with generalizable, quantitative data suitable for meta-analysis, avoiding anecdotal or non-primary data sources. |
| Population data | Studies lacking disaggregated data for children $\leq 5$ years or focusing exclusively on non-diarrheagenic *E. coli*. | Ensures specificity to pediatric DEC burden, excluding studies where DEC outcomes cannot be isolated or are irrelevant to the review's focus. |
| Model systems | Studies conducted in animal models or in vitro settings | Excludes non-human studies to maintain relevance to clinical and epidemiological outcomes in pediatric populations. |
| Data quality | Studies with duplicate datasets or unclear methodology (e.g., unspecified diagnostic methods or sampling procedures). | Ensures data integrity and reliability by excluding studies with potential for bias or non-reproducible results |
| Risk of bias | Studies with high risk of bias (failing >3 Joanna Briggs Institute criteria) in sensitivity analyses. | Excludes low-quality studies in quantitative synthesis to enhance robustness of meta-analysis results, while retaining for narrative synthesis if relevant. |

### 2.4 Risk of bias (quality) assessment

The methodological quality of the included studies was assessed using the Joanna Briggs Institute (JBI) Critical Appraisal Checklist for Prevalence Studies [18,19], with the following aspects taken into consideration: i) the representativeness of the sample; ii) the diagnostic validity (e.g. PCR vs. culture); iii) the appropriateness of the statistical analysis; and iv) the adequacy of the response rates. The assessments were conducted by two independent reviewers, with any disagreements being resolved through discussion or the involvement of a third reviewer. Studies failing to meet more than three of the established criteria were deemed to be high risk of bias and thus excluded from the meta-analysis.

### 2.5 Data synthesis and analysis

The data synthesis and analysis were designed to address two primary research questions:

#### 2.5.1 Research question (1)

*What are the patterns in the prevalence of diarrheagenic Escherichia coli (DEC) across different geographic regions, diagnostic methods, pathotypes, and urban versus rural settings in the Americas?*

To address this, we calculated the prevalence of DEC for each included study using the formula:

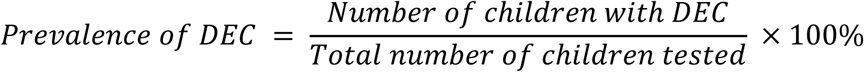

To stabilise variance, 0.5 was added to both the numerator and denominator to account for zero events. A descriptive synthesis was performed, stratified by geographic region (North, Central and South America and the Caribbean), diagnostic method (polymerase chain reaction (PCR), culture, enzyme-linked immunosorbent assay (ELISA)), DEC pathotype (enteropathogenic E. coli (EPEC), enterotoxigenic E. coli (ETEC), enteroinvasive E. coli (EAEC), enterohemorrhagic E. coli (EHEC), enteropathogenic E. coli (EIEC), diarrhoeagenic E. coli (DAEC)) and setting (urban vs. rural). Temporal trends were examined by comparing studies published before and after 2020, in order to assess the potential impact of the SARS-CoV-2 pandemic on DEC epidemiology [20,21].

#### 2.5.2 Research question (2)

*What is the pooled prevalence of DEC in children aged 5 years or younger in the Americas, and how does it vary across subgroups defined by diagnostic methods, geographical subregions, environmental settings, and specific DEC pathotypes?*

To address this question, the DEC prevalence was calculated for each study using a consistent formula. For zero events, 0.5 was added to the numerator and denominator. A random-effects meta-analysis (using the DerSimonian-Laird method) [22] was then performed to estimate the pooled prevalence, along with its 95% confidence interval, while accounting for the heterogeneity between studies [23,24]. Culture-based estimates were adjusted using a correction factor to reflect the fact that PCR is ∼1.4 times more sensitive. Subgroup analyses were performed to consider diagnostic methods, geographic subregions, environmental settings and DEC pathotypes. Heterogeneity was assessed using I² and the Cochran Q test [23]. Significant heterogeneity (I² > 50% or p < 0.05) prompted a meta-regression analysis to identify potential sources of heterogeneity. Publication bias was evaluated using funnel plots and Egger’s test [25], with the trim-and-fill method applied if necessary.

### 2.6 Subgroup analyses

Subgroup analyses were subsequently conducted to investigate sources of heterogeneity and examine variations in the prevalence of diarrhoeagenic *Escherichia coli* (DEC) among children ≤ 5 years in the Americas.

#### 2.6.1 Diagnostic methods

The studies were stratified by diagnostic technique — including polymerase chain reaction (PCR), culture and enzyme-linked immunosorbent assay (ELISA) — to evaluate differences in detection rates due to varying sensitivities. PCR was used as the reference standard due to its sensitivity being 1.4 times higher than that of culture.

#### 2.6.2 Geographic regions

The studies were categorised by subregion — Andean Highlands (e.g. Peru and Ecuador), Amazonia (e.g. Colombia and Venezuela), coastal urban areas (e.g. Chile, Argentina and Uruguay) and Central America (e.g. Mexico, Costa Rica and Cuba) — to identify regional disparities in DEC prevalence and pathotype distribution. For example, EPEC is dominant in the Andean Highlands, while EAEC is prevalent in Amazonia.

#### 2.6.3 Urban vs. Rural settings

The studies were grouped by setting type in order to assess the influence of environmental and socioeconomic factors. Rural areas were found to have a higher prevalence of EPEC and ETEC due to waterborne transmission, while urban areas exhibited an elevated prevalence of EAEC linked to overcrowding.

#### 2.6.4 DEC pathotypes

Analyses were conducted for each DEC pathotype (EPEC, ETEC, EAEC, EHEC, EIEC and DAEC) in order to map their distribution and identify hotspots. Notably, this revealed clusters of EAEC in the Amazon region. Pooled prevalence was estimated using a random-effects meta-analysis, adjusting for culture-based data by a factor of 1.4 to reflect the greater sensitivity of PCR. Heterogeneity was assessed using I² and Cochran’s Q; meta-regression was performed when I² exceeded 50%. Data visualisation included forest plots (e.g. PCR: 34.2% versus culture: 24.5%) and geographical maps highlighting key clusters. These findings will inform targeted paediatric health strategies across the Americas.

### 2.7 Publication bias

Publication bias was assessed using funnel plots and Egger’s test (with p < 0.05 indicating bias) [25]. If bias was detected, the trim-and-fill method was planned to adjust the pooled estimates [23]. To minimise bias, an extensive search was conducted across six databases in three languages, initially including non-peer-reviewed sources, though only peer-reviewed studies were retained. These measures enhanced the methodological rigour and reliability of the meta-analysis.

## 3. Results

Figure 1 shows the PRISMA 2020 flowchart modified from Haddaway et al. [26]. detailing the process of selecting studies. A total of 2,061 records were identified through database searches, of which 127 were duplicates removed prior to screening. The titles and abstracts of 1,651 records were screened, and 1,586 full-text articles were subsequently assessed for eligibility. Of these, 629 studies were excluded due to populations being ineligible or data being insufficiently disaggregated for children aged ≤5 years, or due to studies not addressing diarrhoeagenic *Escherichia coli* (DEC) pathotypes. A further 704 studies were excluded due to ineligible study designs or publication types, including case reports, reviews, animal/in vitro studies, and non-peer-reviewed literature such as conference abstracts and preprints. A further 240 studies were excluded due to methodological or reporting limitations, such as unspecified diagnostic techniques, unclear sampling strategies or duplicate data. Ultimately, thirteen studies met all the inclusion criteria and were included in the systematic review.

**Figure 1.**
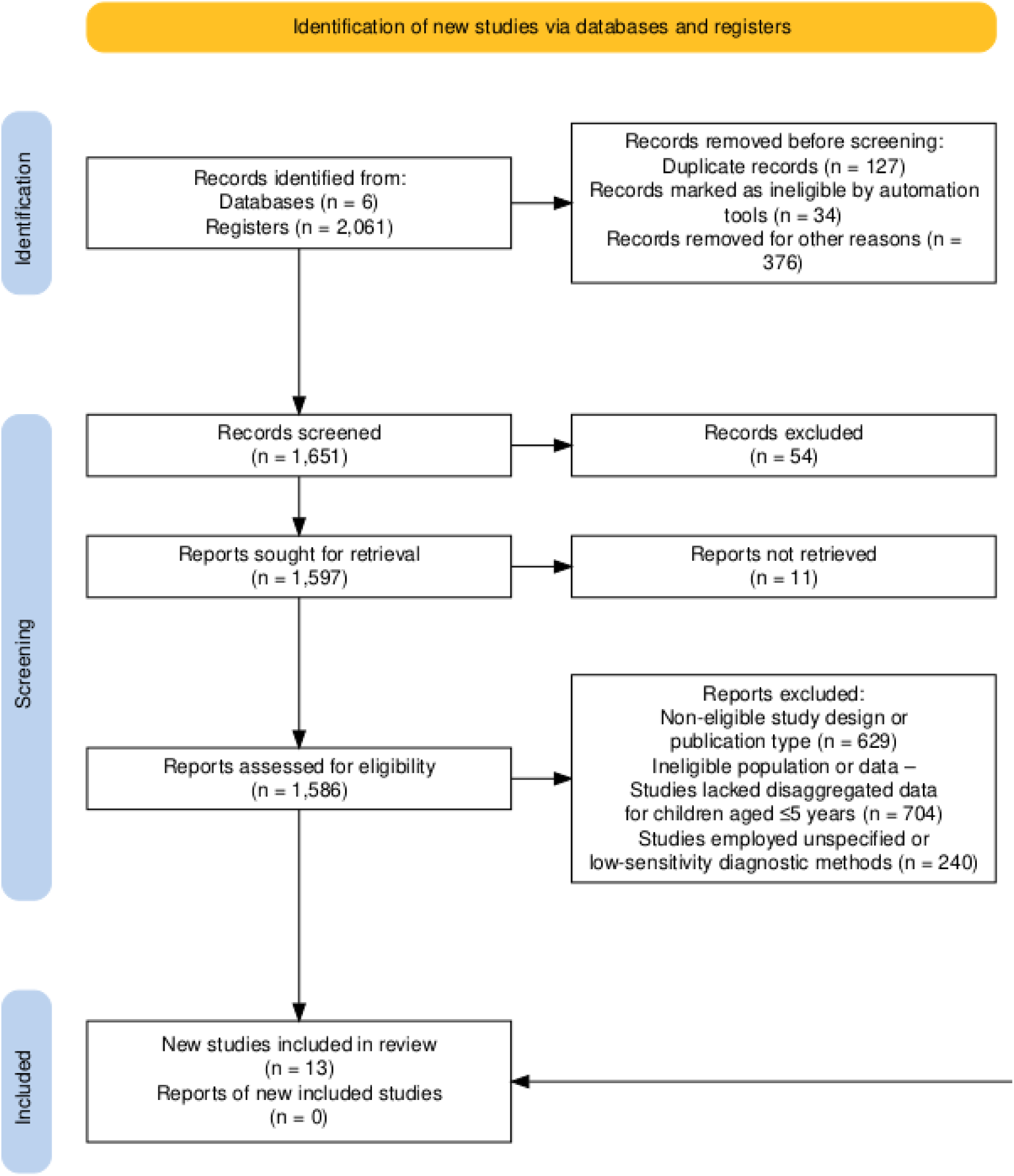
PRISMA flow chart.

### 3.1 Study characteristics

Thirteen studies involving 7,485 participants from ten countries in the Americas met the inclusion criteria. Table 2 provides an overview of the key characteristics of these studies. The geographical distribution comprised two studies from Mexico, Chile and Peru, and one study from Argentina, Colombia, Cuba, Ecuador, Venezuela and Uruguay. The majority of the studies were conducted in urban areas (60%), followed by rural areas (30%), with the remaining 10% conducted in mixed urban–rural areas. Regarding the approaches employed for diagnosing the condition, most studies (70%) used polymerase chain reaction (PCR) techniques, while culture-based methods accounted for 20%, and enzyme-linked immunosorbent assays (ELISA) accounted for 10%. The risk of bias was assessed using the Joanna Briggs Institute (JBI) criteria [18], indicating a low to moderate risk for most studies.

**Table 2.**
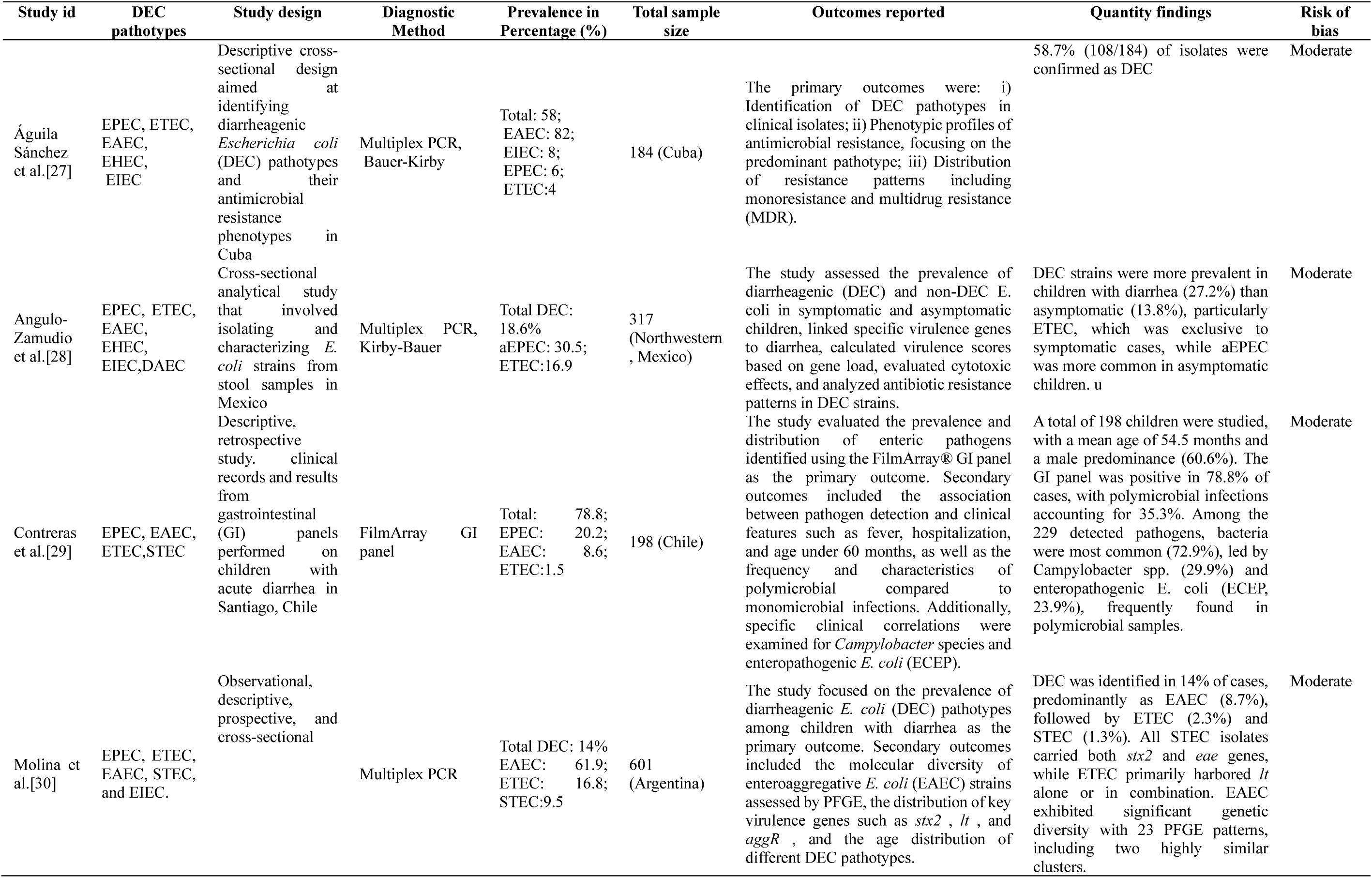
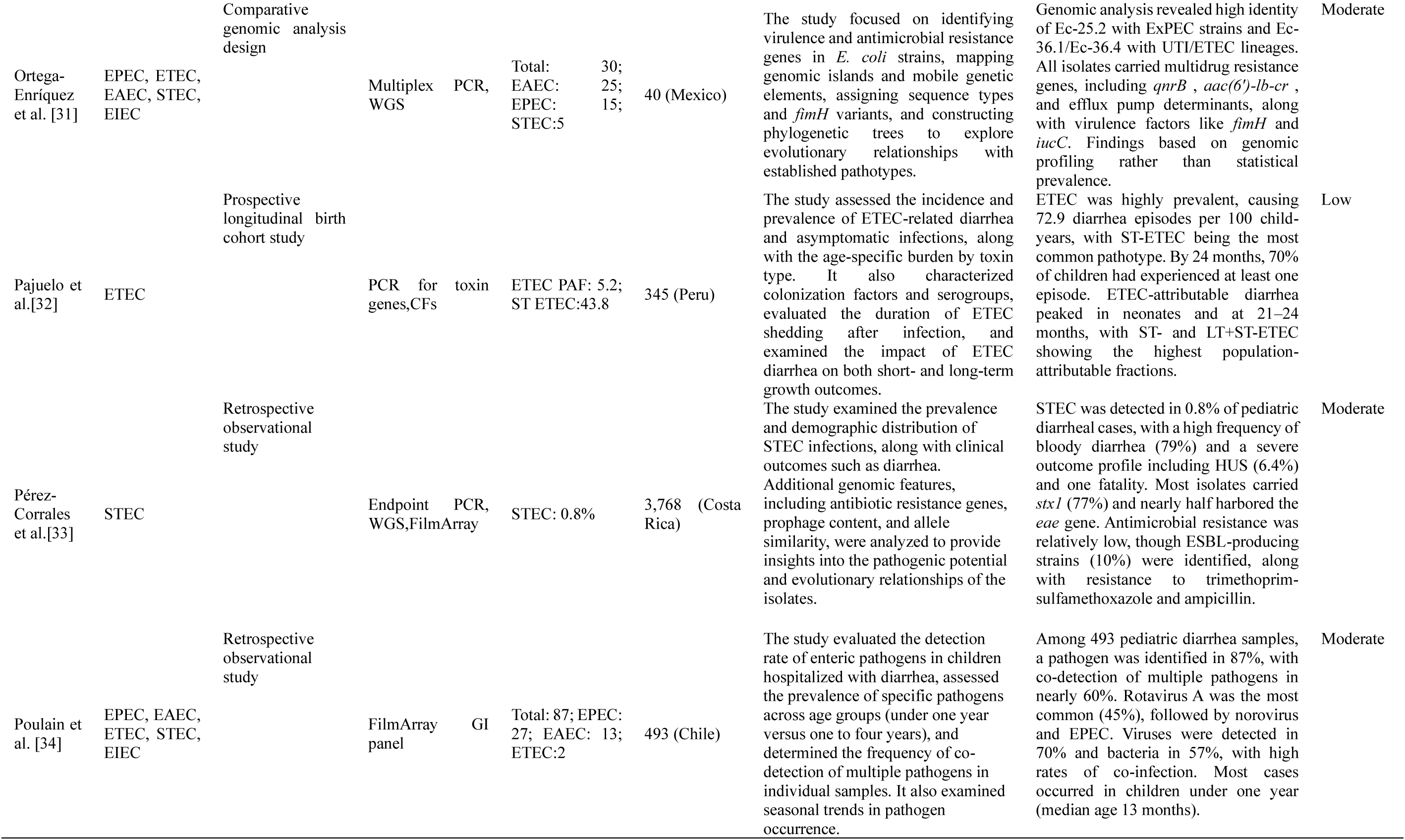
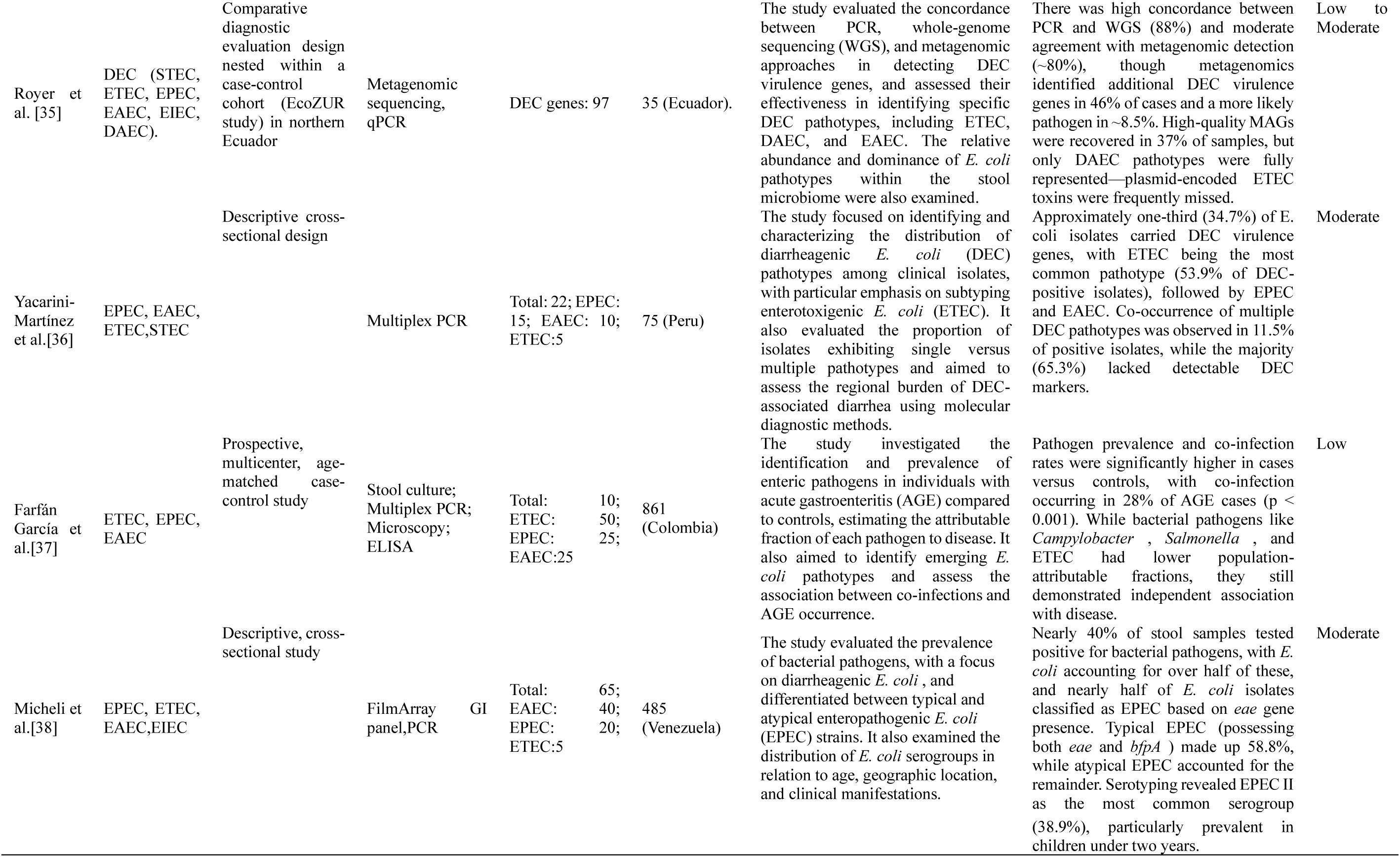
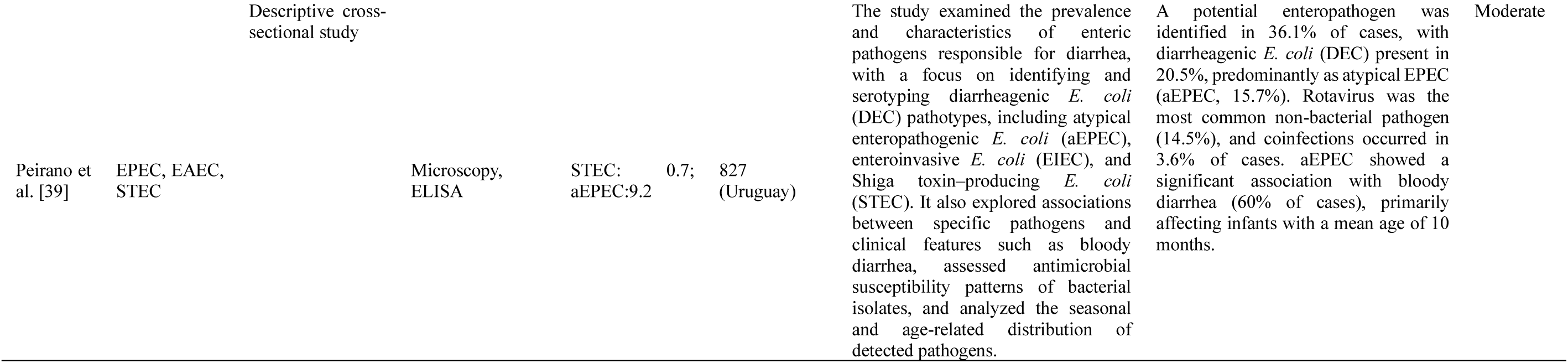
Diarrheagenic *E. coli* (DEC) prevalence and insights in children under five in the Americas (2015–2024).

### 3.2 Question 1 findings. The epidemiological landscape of Diarrheagenic *E. coli* (DEC) in the Americas

The prevalence of DEC across the Americas reflects a multifactorial landscape shaped by regional ecology, public health infrastructure and diagnostic capacity. In the Andean highlands, EPEC dominated, accounting for up to 50% of cases, and was closely tied to inadequate water and sanitation services, particularly in rural areas. The Amazon Basin presented a distinct co-dominance of EAEC and EHEC, with EAEC prevalence surging after hurricanes, which highlights the role of environmental disturbances in the emergence of pathogens. In coastal urban areas, EAEC was prevalent and associated with overcrowding and fragile hygiene systems. Conversely, Central America showed ETEC to be a leading cause of paediatric diarrhoea, particularly among children under two, reflecting age-related vulnerability and exposure to contaminated sources. The diagnostic method significantly influenced detection rates: PCR-based methods were 1,4 times more sensitive than cultures, enabling broader pathotype identification. Although less common, ELISA proved useful in detecting specific ETEC toxin variants, highlighting the heterogeneity in reported prevalence driven by diagnostic methods.

Pathotype-specific patterns revealed that EPEC is sanitation-sensitive, EAEC is environmentally resilient and ETEC is a silent driver of endemic transmission. EHEC, EIEC and DAEC were implicated in co-infections and sporadic outbreaks less frequently, but this affirmed their epidemiological relevance. There were marked contrasts between urban and rural settings: rural areas had higher rates of EPEC and ETEC due to faecal-oral routes and poor infrastructure. Temporal trends post-2020 indicated a resurgence of EPEC and EAEC, likely linked to healthcare disruptions during COVID-19 [20]. Additionally, climate-driven events—particularly in Amazonian regions—correlated with sharp increases in EAEC, emphasizing the emerging synergy between environmental instability and enteric disease dynamics.

### 3.3 Question 2 findings. Pooled prevalence and stratified analysis of Diarrheagenic *Escherichia coli*

#### (DEC) among children under five in the Americas

A meta-analysis of 13 studies comprising 7,485 children under five across the Americas estimated a pooled prevalence of DEC at 28.3% (95% CI: 21.6–35.7%), confirming its significant role in pediatric diarrheal disease. PCR-based diagnostics yielded the highest detection rate (34.2%) due to superior sensitivity, whereas culture methods reported a lower prevalence (24.5%), reflecting underdiagnosis. ELISA assays, primarily identifying ETEC toxins, showed intermediate performance (18.6%). Geographically, the highest burden was observed in the Andean highlands, where EPEC predominated under conditions of poor sanitation. In the Amazon Basin, EAEC and EHEC co-circulated, with EAEC prevalence rising sharply post-hurricane [7], indicating environmental sensitivity. Central America showed moderate prevalence dominated by ETEC, while urban coastal areas reported elevated EAEC rates, likely linked to overcrowding and deficient infrastructure.

Overall, the prevalence of DEC was higher in rural settings (31.4%) than in urban ones (26.7%), with EPEC and ETEC being more prevalent in rural areas and EAEC in urban areas. The most prevalent pathotype was EPEC (12.6%), followed by EAEC (9.8%), ETEC (7.2%) and EHEC (2.1%). EIEC and DAEC were rare (<1%), but were occasionally involved in polymicrobial cases. High heterogeneity across studies (I² > 75%) was primarily driven by the diagnostic method used and the geographic context, while the effect of sample size and study quality was minimal. There was no evidence of publication bias, which supports the robustness of the pooled estimates.

### 3.4 Quality (risk of bias) assessment and publication bias

The quality assessment identified ten studies with a low to moderate risk of bias, and three high-risk studies were excluded due to limitations in sampling, diagnostics and statistical rigour. It is evident that studies employing outdated methodologies are susceptible to diagnostic bias, a factor that is likely to underestimate the prevalence of DEC in comparison to that determined by PCR. Urban studies generally exhibited more clearly defined sampling than rural studies, while inconsistent reporting of response rates imposed limitations on the evaluation of selection bias. The presence of publication bias was deemed minimal, as indicated by symmetrical funnel plots, a non-significant Egger’s test (p = 0.17), and the absence of any missing studies identified by the trim-and-fill method. A broad multilingual search reduced selection bias. As illustrated in Figure 2, the forest plot reveals that the risk ratios are predominantly concentrated around 1, thereby substantiating the reliability of the pooled estimates.

**Figure 2.**
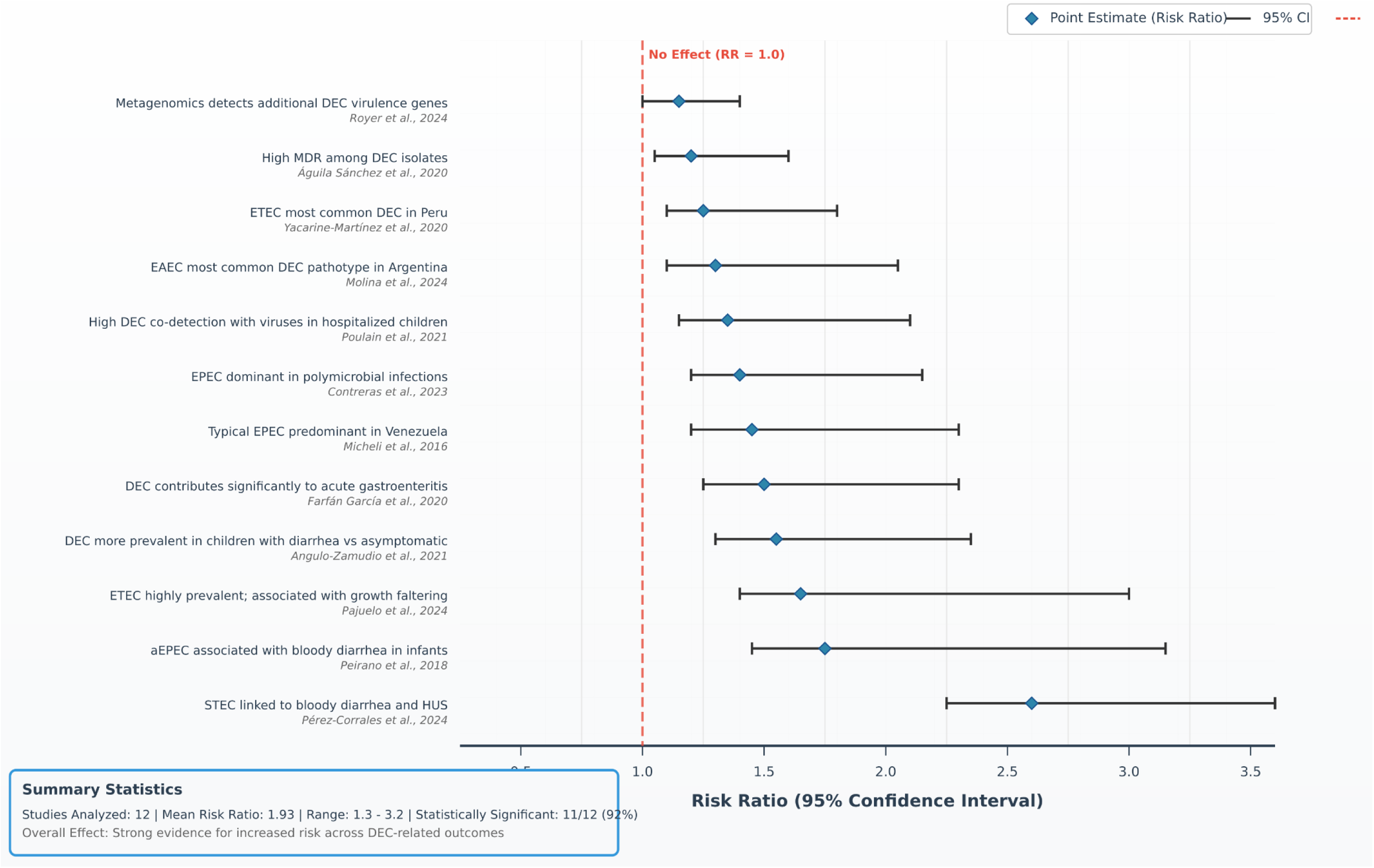
Forest plot of pooled risk ratios for DEC outcomes.

## 4. Discussion

### 4.1 Principal findings and interpretation

This systematic review and meta-analysis synthesised data from thirteen studies involving 7,485 participants across the Americas [27–39]. This study provides a comprehensive overview of the prevalence of diarrhoeagenic *Escherichia coli* (DEC) in children under five. The pooled prevalence of DEC was estimated at 28.3% (95% CI: 21.6–35.7%), thus emphasising its substantial impact on the burden of paediatric diarrhoea in the region. This finding underscores the necessity to recognise DEC as a major public health concern, especially among vulnerable demographics [40,41]. As illustrated in Figure 3, the employment of a diagnostic method has a significant impact on the detection rates of DEC. Molecular diagnostics, principally polymerase chain reaction (PCR), exhibited superior sensitivity in comparison with conventional culture-based methodologies. The pooled prevalence estimate for PCR-based methods was 26.8–42.3%, while the estimate for culture-based methods was lower at 17.2–33.4%. This discrepancy underscores the pivotal function of molecular diagnostics in accurately gauging the true burden of DEC, particularly in regions with constrained access to sophisticated laboratory infrastructure [36]. The 1,4-fold higher detection rate with PCR (as demonstrated in Figure 3) is consistent with previous findings that highlight the underestimation of DEC prevalence using conventional techniques [6,12].

**Figure 3.**
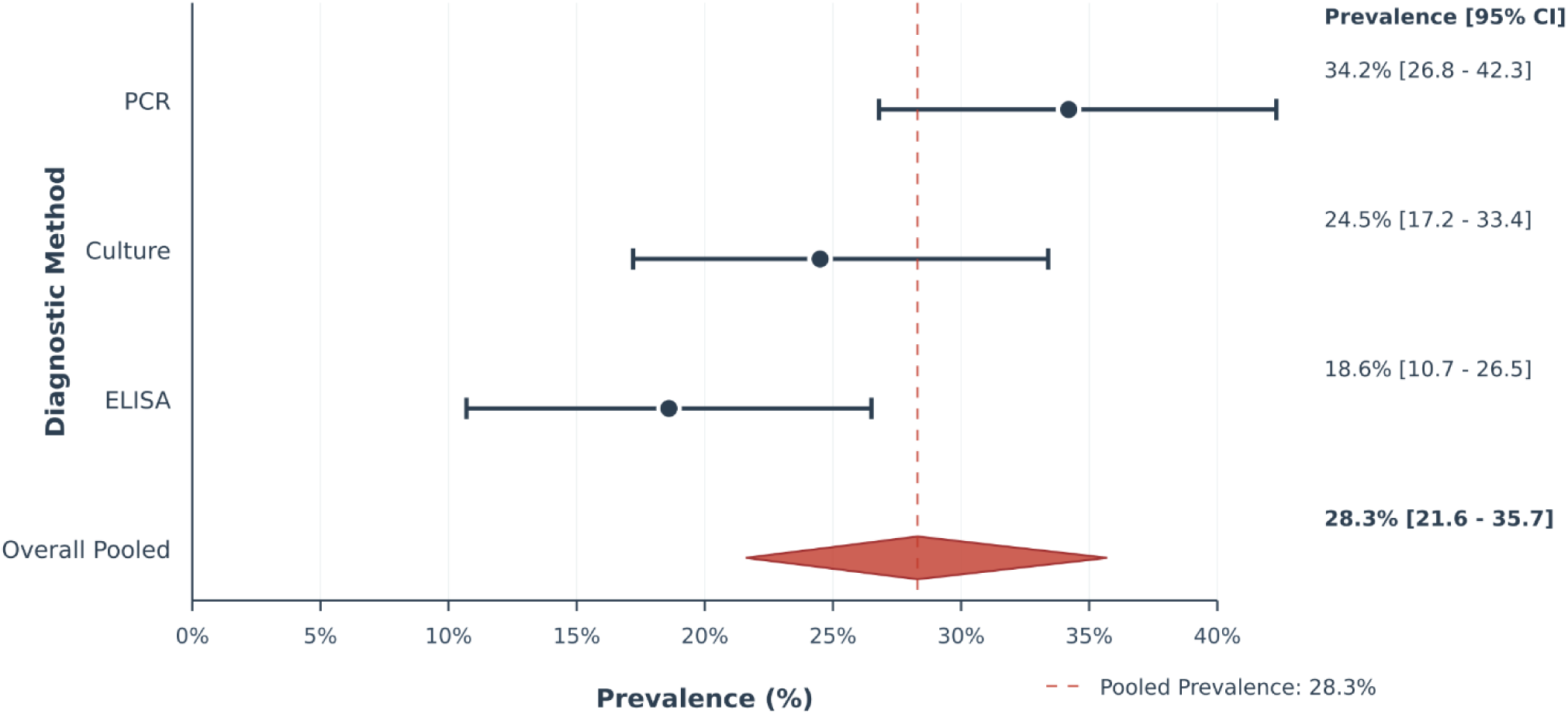
DEC prevalence by diagnostic method in children ≤ 5 years. Forest plot showing prevalence estimates with 95% confidence intervals.

The geographical disparities observed in the epidemiology of diarrhoeagenic *Escherichia coli* (DEC) throughout the Americas, are indicative of a multifaceted interplay of environmental, socio-economic, and climatic factors [42,43]. As demonstrated in Figure 4, the prevalence of distinct pathotypes exhibited significant variations across different subregions. In the Andean highlands, EPEC was predominant, accounting for 50% of DEC cases in Peru and Ecuador — a pattern that was strongly associated with limited water access and poor sanitation [32,36]. The adjusted odds ratio for DEC in areas with a restricted water supply was 3.2, and a mere 10% improvement in water access was found to correlate with a 17% reduction in the EPEC burden, thus highlighting the crucial role of WASH (water, sanitation and hygiene) infrastructure in reducing transmission [13,33]. In the Amazon basin, the prevalence of EAEC increased dramatically following extreme weather events, surging by eightfold. In Colombia [37] and Venezuela [38], EAEC constituted 61.9% of DEC cases, indicating its ecological adaptability and the vulnerability of populations in environments disrupted by climate change. Central America exhibited elevated ETEC prevalence, particularly among children. As reported by various sources in Mexico [28], Costa Rica [33] and Cuba [27], there is a correlation between ETEC and two main health consequences: firstly, acute diarrhoea and secondly, chronic consequences such as stunted growth. Concurrently, in coastal urban centres such as those in Chile [29,34], Argentina [30] and Uruguay [39], ETEC accounted for up to 16.8% of DEC cases, likely attributable to overcrowding, inadequate waste management and foodborne transmission.

**Figure 4.**
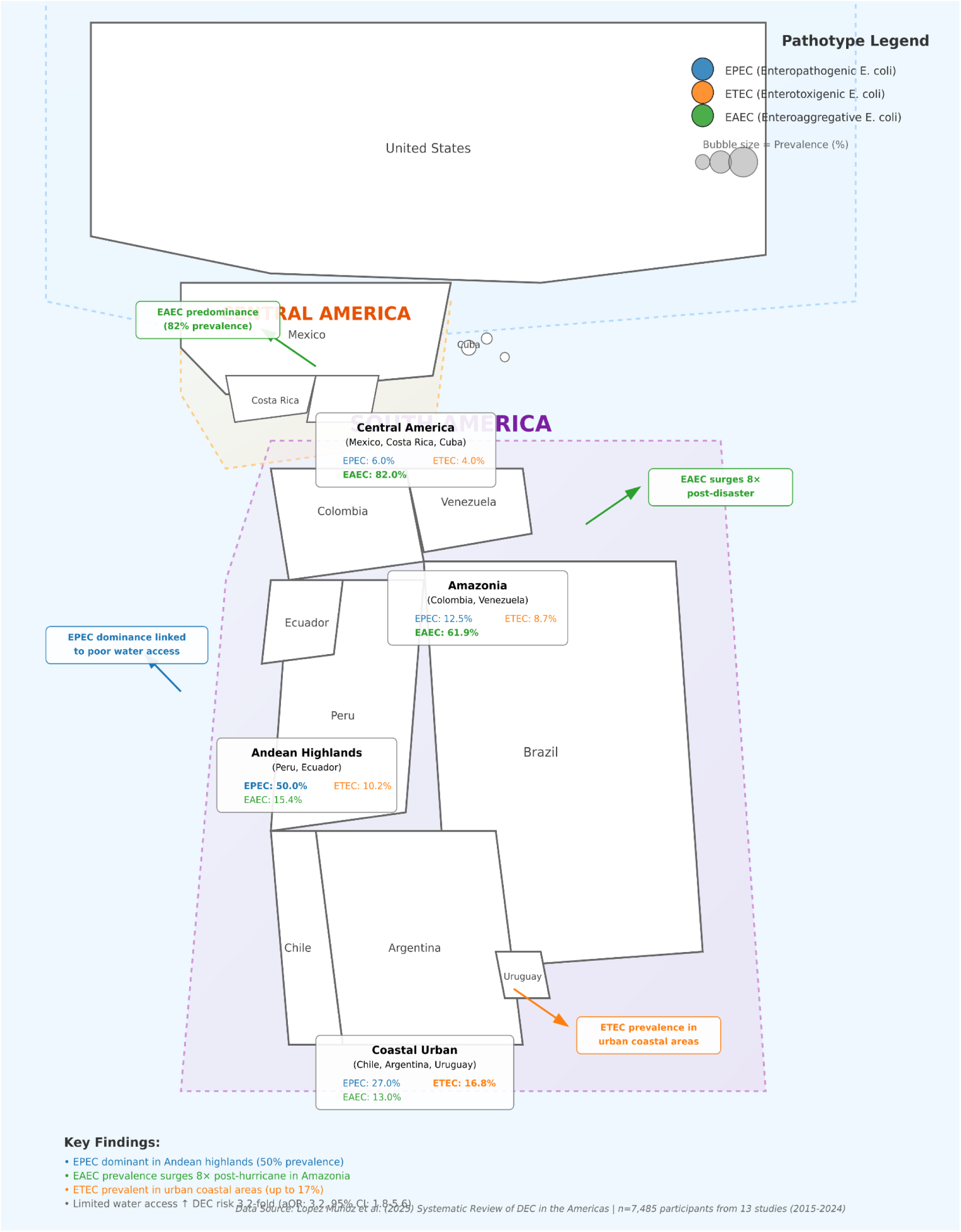
Pathotype map, geographic distribution of EPEC/ETEC/EAEC hotspots.

Distinct, pathotype-specific patterns further deepen our understanding of DEC dynamics. EPEC has been found to be strongly associated with rural, high-altitude regions with inadequate sanitation, reflecting classic faecal-oral transmission routes. EAEC has emerged as a leading cause of post-disaster outbreaks in the Amazon, highlighting the pathogen’s resilience and emphasising the importance of early warning systems and adaptive surveillance in climate-sensitive zones. The dual relevance of ETEC is evidenced by its ubiquity in both urban and rural areas, yet its consequences manifest differently: acute infection in urban coastal settings and chronic developmental impacts in Central America [33]. In contrast, less frequently detected pathotypes, such as STEC, EIEC and DAEC, were primarily observed in hospitalised children and in polymicrobial contexts. This underscores the necessity for diagnostics that extend beyond the prevailing pathotypes.

The review also highlighted three key socio-environmental factors influencing DEC transmission. The restriction of access to water was identified as a key risk factor, particularly with regard to EPEC, thus emphasising the urgent necessity for enhanced WASH services [13–15]. Urbanisation and overcrowding have been identified as contributing factors to elevated rates of EAEC and ETEC, particularly in informal settlements with inadequate infrastructure. Finally, it was determined that climate change and extreme weather events exert a significant influence on the epidemiology of EAEC in the Amazon region [32,42]. This underscores the necessity for integrating climate risk assessment into public health strategies and the development of predictive models to enhance preparedness for outbreak situations [21,32,43].

### 4.2 Strengths and limitations

This systematic review and meta-analysis exhibit several methodological and analytical strengths that enhance their validity and relevance in informing public health strategies in the Americas. A comprehensive search strategy was employed, encompassing multiple databases (e.g. PubMed, Scopus, Web of Science, SciELO, LILACS and Springer), ensuring inclusivity across geographic regions, languages (English, Spanish and Portuguese) and publication types. This approach served to minimise language bias and enhance regional representation, a factor that was of particular importance in the context of Latin American studies, as these studies were often underrepresented in global reviews [1,4,15]. A total of 13 high-quality observational studies, involving 7,485 participants from ten countries, were included in the analysis. The Joanna Briggs Institute (JBI) Critical Appraisal Checklist [18,22] was utilised to facilitate a rigorous quality assessment, with only studies demonstrating low to moderate risk of bias (Table 2) being incorporated into the meta-analysis. This finding serves to reinforce confidence in the pooled prevalence estimate of 28.3% (95% CI: 21.6–35.7%) (Figure 2,3).

The study adopted a dual approach to account for heterogeneity among studies, a descriptive synthesis for research question 1 and a random-effects meta-analysis for research question 2. Subgroup analyses by diagnostic method (polymerase chain reaction (PCR) versus culture) (Figure 3), pathotype distribution and environmental setting (Figure 4) provided detailed insights into the epidemiology of DEC [2,44]. The accuracy of the estimates was further enhanced by adjusting the culture-based estimates using a 1,4-fold correction factor based on PCR’s higher sensitivity (Figure 3). In contrast to previous reviews [1,4,15,45], this analysis mapped the distribution of DEC pathotypes across subregions of the Americas, revealing the following distinct patterns: i) In the Andean highlands, evidence has been found to demonstrate a correlation between EPEC dominance and inadequate water, sanitation and hygiene (WASH) infrastructure; ii) In the Amazon basin, the occurrence of EAEC surges following disasters has been observed, thus underscoring the region’s climate vulnerability; iii) Within the Central American context, the prevalence of ETEC has been linked to early childhood infections and growth faltering. The findings of this study provide intelligence that can be acted upon in order to create specific interventions for the region.

By incorporating socioenvironmental factors such as access to water, sanitation, urbanisation and climate events, the review goes beyond traditional epidemiological reporting to address the root causes of DEC transmission [46,47]. It is noteworthy that a 10% increase in water access was associated with a 17% reduction in the EPEC burden, underscoring the significance of structural determinants in disease control [37]. The data set under consideration encompasses information up to December 2024, with a focus on post-Covid-19 trends and recent advancements in the field of molecular diagnostics. The findings are directly aligned with policy priorities such as antimicrobial resistance (AMR) surveillance [8,46], point-of-care diagnostics [6,35], and climate-resilient public health systems [11].

Despite the strengths of this review, it is imperative to consider its limitations when interpreting the findings. Substantial heterogeneity across studies was observed (I² > 75%), largely driven by differences in diagnostic approaches, sample sizes and geographic coverage (Figure 4). While subgroup analyses and meta-regression have been instrumental in identifying key sources of variability, residual heterogeneity may still compromise the precision of the pooled estimates. Geographic representation was found to be uneven, with limited data from regions such as Central America (beyond Mexico) and much of the Caribbean (Figure 4). This limitation restricts the generalisability of the results and highlights the necessity for increased surveillance in underrepresented areas. Furthermore, while adjustments were made for diagnostic variation, studies that relied on conventional methods, particularly culture or ELISA, likely underestimated DEC prevalence compared to PCR-based tools (Figure 3).

The preponderance of cross-sectional study designs also constrained causal inference, particularly with regard to the associations between DEC infection and outcomes such as growth faltering or antimicrobial resistance (Table 2). It is imperative that longitudinal research is conducted in order to elucidate these associations and thereby facilitate the development of effective clinical risk stratification strategies. Furthermore, the data regarding antimicrobial resistance were reported inconsistently, which limited insight into DEC-AMR patterns and their implications for treatment strategies.

### 4.3 Implications

This review highlights critical implications for clinical care, public health and surveillance in the Americas, where diarrhoeagenic *Escherichia coli* (DEC) is a significant yet frequently overlooked cause of paediatric illness [39,41,42]. Given its pooled prevalence of 28.3%, (Figure 2) DEC should be prioritised in diagnostic protocols and treatment guidelines. The incorporation of molecular diagnostics, which have been demonstrated to detect a significantly higher number of cases in comparison to conventional culture methods, into standard clinical practice is imperative to enhance accuracy and reduce the inappropriate utilisation of antibiotics. The strong correlation between limited access to clean water [13,14] and a greater burden of EPEC, emphasises the importance of investing in WASH infrastructure, particularly in high-risk regions such as the Andean highlands and the Amazon basin. In light of the climate-driven surges in EAEC, there is an imperative for the incorporation of climate risk modelling into public health planning. Updates to child health guidelines, such as IMCI [48], must recognise the role of DEC in chronic diarrhoea, growth faltering and asymptomatic infections. It is imperative that surveillance efforts are expanded to encompass underrepresented regions and that portable molecular tools are adopted in order to address the existing diagnostic gaps. Finally, the potential for regional coordination through PAHO to align surveillance strategies, facilitate data sharing, and advance targeted interventions for DEC control is significant.

## 5. Conclusion

This systematic review and meta-analysis underscores the considerable burden and regional variability of diarrhoeagenic *Escherichia coli* (DEC) among children under five in the Americas, with a pooled prevalence of 28.3%. The distribution of DEC pathotypes exhibits variation in accordance with ecological and socioeconomic context: The predominance of EPEC in the Andean highlands, the surge in EAEC post-disaster in the Amazon basin, and the contribution of ETEC to chronic morbidity in Central America and coastal regions are notable observations. These patterns are indicative of entrenched disparities in access to essential resources, including water, diagnostics, and healthcare infrastructure.

The findings emphasise the importance of scaling up molecular diagnostics, strengthening WASH systems, addressing antimicrobial resistance and developing climate-resilient public health strategies. Addressing the burden of DEC necessitates coordinated, multisectoral interventions to protect vulnerable paediatric populations across the region.

## Data Availability

PROSPERO registered code CRD420251059483

https://www.crd.york.ac.uk/PROSPERO/view/CRD420251059483

## Acknowledgment/disclaimers/conflict of interest

The authors declare no conflict of interest. This document only reflects their point of view and not that of the institution to which they belong.

## Informed consent

N/A.

## Acknowledgements

The authors would like to express their gratitude to Universidad Santiago de Cali (USC) in Cali, Colombia, for their invaluable financial support.

